# Development of a Codebook of Online Anti-Vaccination Rhetoric to Manage COVID-19 Vaccine Misinformation

**DOI:** 10.1101/2021.03.23.21253727

**Authors:** B. Hughes, C. Miller-Idriss, R. Piltch-Loeb, K. White, M. Creizis, Caleb Cain, E. Savoia

**Author notes:** **Authors’ contribution**: Dr. Hughes oversaw and participated in codebook data sampling, code development, analysis of coding data, and final code selection, as well as writing descriptive sections of codebook, selecting example images, and drafting of manuscript. Dr. Hughes contributed to the survey instrument and provided feedback during the development of the manuscript. Dr. Miller-Idriss contributed to the study design and writing of the findings. Dr. Savoia and Dr. Plitch-Loeb contributed to the interpretation of findings and development of the manuscript.

## Abstract

Vaccine hesitancy (delay in obtaining a vaccine, despite availability) represents a significant hurdle to managing the COVID-19 pandemic. Vaccine hesitancy is in part related to the prevalence of anti-vaccine misinformation and disinformation, which are spread through social media and user-generated content platforms. This study uses qualitative coding methodology to identify salient narratives and rhetorical styles common to anti-vaccine and COVID-denialist media. It organizes these narratives and rhetorics according to theme, imagined antagonist, and frequency. Most frequent were narratives centered on “corrupt elites” and rhetorics appealing to the vulnerability of children. The identification of these narratives and rhetorics may assist in developing effective public health messaging campaigns, since narrative and emotion have demonstrated persuasive effectiveness in other public health communication settings.

## Introduction

The COVID-19 pandemic has arisen in a period of increasing vaccine hesitancy in the United States. Vaccine hesitancy is defined as a delay in the acceptance of a vaccine, or the outright refusal to take a vaccine despite its availability (Lahouati, 2020). Vaccine hesitancy is of particular concern when a vaccine is the primary method to mitigate the spread of a serious disease. Prior to the beginning of the COVID-19 pandemic, vaccine hesitancy was declared one of the top ten threats to global health by the World Health Organization (World Health Organization, n.d.). The pandemic emerged shortly after a 2019 measles outbreak which has since been tied to parental reluctance to vaccinate schoolchildren (Benecke & DeYoung, 2019; Olive et al., 2018). This was the worst outbreak of its kind since 1992 (CDC, 2021) and since a historic low of 0.15 measles cases per million in 2002 (CDC, 2021) outbreak.

Several national opinion polls have found a significant portion of the US population is hesitant to take a COVID-19 vaccine. The prevalence of COVID-19 vaccine hesitancy has ranged from a quarter to half of the US population depending on the time in which surveys were conducted, reflecting the ongoing challenge of addressing vaccine hesitancy in the pursuit of herd immunity ((Muñana et al., 2020); Ipsos, 2020; Cornwall, 2020; Johnson, 2020;).

### The Role of Online Fora and Social Media

Multiple mechanisms can contribute to the spread of vaccine hesitancy, among which online forums play an important role in persuasion (Dubé et al., 2013; Goldstein et al., 2015; Obregon et al., 2009). In the past, persuasion might have occurred via a “two-step flow,” in which opinion leaders were integral to the adoption of people’s worldviews (Katz & Lazarsfeld 1955). However, with the growth of increasingly personalized and automatically customized content channels offered on the web, some scholars have observed the emergence of a complementary “one-step flow” of persuasion (Bennet, Manheim 2006; Thorson, Wells 2016). In this “one-step flow,” web users are influenced directly from an online experience that has been algorithmically tailored to address their interests—and psychological vulnerabilities (Nadler, Crain, & Donovan 2018). The present study is informed by the hybrid theory of Hilbert et. al (2017), which argues that while persuasion has indeed become less interpersonal, and more automated and direct, online influencers and trusted offline sources still play a crucial role in shaping opinion.

Several authors have examined how social media platforms contributed to vaccine hesitancy prior to the COVID-19 outbreak (Basch et al 2019; Gunarante et al 2019; Arif et al, 2018; Moran 2016; Ekram 2019). Amplification of vaccine hesitancy online has continued in light of development of the COVID-19 vaccine (Puri 2020). Studies reviewed by Puri and colleagues (2020) describe how anti-vaccine content frequently generates greater user engagement than its pro-vaccine counterparts on Facebook. Singh and colleagues (2020) found that low quality sources of misinformation on COVID-19 were more commonly retweeted than those with high quality information. In a comparative analysis of the spread of misinformation on five social media platforms (Twitter, Instagram, YouTube, Reddit and Gab), Cinelli and colleagues (2020) analyzed more than 8 million comments and posts over a time span of 45 days, to model the spread of misinformation and demonstrated that social media platforms can serve as amplifiers of misinformation.

Vaccine hesitancy at large often arises in response to real world events that create a window of opportunity for narratives countering vaccine uptake. According to Betsch et. al, even limited and short-term exposure to anti-vaccine websites increased individual perceptions of vaccine risks (Betsch et al., 2010). Buller and colleagues (2019) have found that an individual’s degree of engagement with Facebook posts on the HPV vaccine was predictive of greater vaccine hesitancy. Buller et. al hypothesize that on the social media platform, alleged risks associated with vaccines, appear more immediate and tangible than risks associated with not getting vaccinated (the success of vaccination is the absence of disease) (Puri, 2020). Further, anti-vaccine messaging tends to be more focused on emotions and personal anecdotes with powerful imagery in contrast to the empirical strategies utilized by pro-vaccination literature and platforms. These emotional approaches tend to be more appealing to social media users and are consistent with other content that tends to be shared on social media (Callender, 2016).

### Types of anti-vax narratives and beliefs

Often vaccine hesitancy is treated by policymakers and risk communicators as a uniform belief; people are *either* hesitant to get a vaccine *or* not hesitant. However, a subset of research has identified ways in which vaccine hesitancy is not a uniform belief system, but rather consists of several tropes and narratives that coalesce around common themes. In a review of 480 websites, all of which were promoting vaccine hesitancy, Moran and colleagues (2016) found several key values among those reviewed: choice, freedom, natural/holism, independence/individuality, and religion. These values were expressed in narratives related to topics ranging from life-style norms (references to alternative messaging or other health behaviors), mistrust in communicating authority, vaccine related diseases, and vaccine ingredients counter to religion (pork products for example). The most common value was “choice” appearing on one-third of the sample identified in the study.

Moran was the first to consider values, or the “core” principles that may drive the more specific beliefs held by—or comments made by—individuals. The study by Moran and colleagues adds to an ongoing literature aiming to characterize the type of anti-vaccination content on the internet (Davies, 2002; Wolfe, 2002; Kata, 2010; Bean, 2011). These prior studies have identified a variety of specific comments or beliefs among content producers. Bean (2011) builds on the work of Davies and Wolfe and refers to specific values that emerge from this content: safety and effectiveness, civil liberties, and alternative treatments. More recent work by Johnson et al. (2020) has found that there is a greater number of smaller anti-vaccination groups online relative to pro-vaccination groups, allowing these groups to cover more “surface area” and attract the undecided to anti-vaccination messages. This “surface area” is further expanded when anti-vaccination groups create content focused on more than just vaccination related issues, connecting to the other core beliefs these authors have described. Critically, John et al. note that anti-vaccine messages tend to be framed as conversations between equals, that is, as peer-to-peer communication. Conversely, pro-vaccine content tends to take the form of expert-to-masses, top-down messages. Anti-vaccine communication thus bridges the gap between one-step and two-step flows of persuasion, perhaps the better to take advantage of online media’s unique style of social ambiguity.

### Defining Rhetoric and Narrative in the Context of Ant-Vaccine and COVID-Denial Content

Whereas these prior studies analyzed the quality of information related to vaccine hesitancy and the values that motivate hesitancy, the study we conducted focuses on the narrative tropes and rhetorical styles of anti-vaccine content. Narratives combined with particular rhetorical strategies can be more or less influential. This study adopts a simple definition of the term *narrative* predicated on the features of change and temporality. Dahlstrom provides an excellent definition of narrative in the context of science communication to mass audiences as “a particular structure that describes the cause-and-effect relationships that take place over a particular time period that impact particular characters” (Dahlstrom, 2014, p. 13614). Our simple definition is in keeping with established scholarly uses of the term (Cuddon, 1999; Genette, 1983; Liveley, 2019; Todorov, 2014).

For the purposes of this study, a *trope* is understood to mean a subsidiary element of narrative, indicative of the larger narrative or narratives in which it is contained (Bahti & Mann, 2012; Burke, 1969; Chandler & Munday, 2011; Cuddon, 1999; Sandberg, 2016)—narratives which might be vague, only partially articulated, or even based on mysterious/absent plot, characters and/or meaning (Foucault, 2012; Nietzsche, 2012; White, 1973). Tropes therefore “are about relationships, and never about the term in itself” (Crocker & Sapir, 2016, p. 3). Tropes, in other words, definitionally refer to something beyond the context in which they appear. A *narrative trope* therefore, is some semiotic element—visual, aural, or written—which connotes a larger story of worldview.

For the purposes of this study, a “rhetorical strategy” refers to those modes of persuasion that can be separated from narrative appeals, which do not in-and-of-themselves indicate a larger story or narrativized worldview. These generally comport with the classical categories of rhetorical appeal: *ethos* (authority), *pathos* (emotion), and *logos* (logic) (Aristotle, 1946) and with the Platonic critique of rhetoric as vulgar and manipulative (Plato, 1842). On one hand, we look to the classical definitions because contemporary definitions of rhetoric are diverse and unresolved (Gunderson, 2009; McNally, 1970; Thomas, 2007; Yoos, 2009) This study takes an expansive view of rhetoric, to include audio-visual as well as written and verbal rhetoric. This is in keeping with common scholarly use of the terms and concept (Finnegan et al., 2008; Handa, 2004; Ommen, 2016; Prelli, 2006). Narrative tropes and rhetorical strategies thus combine to form persuasive messages.

### The Efficacy of Narrative and Rhetorical Persuasion vs. Appeals to Reason

While the literature is not uniform in its conclusions (Hinyard & Kreuter, 2007; Krause & Rucker, 2020), there is evidence to suggest that narrative and perspectival persuasion can be more effective than factual-argumentative approaches (Adebayo et al., 2020; Carrion, 2018; Chang, 2008; de Wit et al., 2008; Krakow et al., 2018; Kreuter et al., 2010; Murphy et al., 2013, 2015; Taylor & Thompson, 1982). Research shows that audience absorption into a narrative reduces audiences’ capacity to form counterarguments against messages contained in the narrative (Deighton & Romer, 1989; Green et al., 2004; Igartua & Barrios, 2012) and tends to strengthen the persuasiveness of weak arguments and appeals (Escalas 2007). As per Roozenbeek and Van der Linden (2020), messages addressing a persuasion technique rather than a specific factual claim offer the potential “to achieve broad-spectrum resistance against manipulation,” which factual rebuttal might not (n.p.).

Braddock’s work in the field of attitudinal inoculation likewise suggests that attention to *form* yields greater negative attitudinal reactance against undesirable belief or behavior than attention to *content* (Braddock, 2019). Ratcliff and Sun argue that narrative’s effectiveness comes from its ability to circumvent psychological resistance, so that “when a persuasive message is embedded in the story and/or carried by the characters, persuasion occurs to the immersed, less critical, and less defensive” (Ratcliff & Sun, 2020, p. 414). And literature from the field of violent extremism studies suggests that individuals are most often brought to false and conspiratorial perspectives not by rational decision-making, but through a search for emotional and social fulfillment (McCauley & Moskalenko, 2016; Miller-Idriss, 2017).

Work in the field of public health messaging has demonstrated that messaging which focuses on narrative and rhetoric (form) tends to yield better persuasive outcomes than messaging which focuses on facts alone (i.e. content) (Adebayo et al., 2020; Bryan et al., 2019; Gilkey et al., 2020; Kreuter et al., 2010; Liu & Yang, 2020; Murphy et al., 2015). A variety of studies from the field of advertising, media, and communication studies suggest that narrative messaging yields more effective outcomes than analytic and informative messages (Bilandzic & Busselle, 2013; Escalas, 2006; Igartua & Barrios, 2012). As a practical matter, Deng et al. found advertising messaging related to COVID-19 deployed “transformational” strategies (that is, appealing to emotions) over informational ones by a ratio of nearly two-to-one (Deng et al., n.d.).

### Study objective

In anticipation of the unfolding COVID-19 vaccination campaign, this study sought to identify narrative tropes and rhetorical strategies in the emerging vaccine hesitancy discourse. To this end, we developed a codebook of the most commonly found anti-vaccination themes, both general and specific to COVID-19. We did so with the intent of providing public health communicators and misinformation managers a codebook of compelling anti-vaccination themes, against which counter-messages might be tailored. This work contributes to the nascent literature on COVID-19 vaccine hesitancy beliefs and the ways in which anti-vaccine messages are persuasive.

## Data Sources and Methods

### Data sources

Data collection aimed to develop corpora (i.e. large collections of codeable content) for two separate rounds of coding. These corpora were developed via purposeful sampling methodology. Using hashtag and keyword searchers, a team of subject matter experts identified 20 channels (i.e. bounded sources of content, such as a social media account) which appeared to contain a high degree of anti-vaccine content and/or COVID-denialism. For each round, five distinct channels were selected for preliminary coding. Only publicly available, English language media were selected, both to comply with platform terms of service and to focus coding efforts on content available to the uncommitted and vaccine hesitant. Geography was not considered, due to the transnational availability of the channels which were surveyed. Channels were selected according to the criteria of 1.) offering 50-100 pieces of codable content from the past six months, 2.) covering written, video, and image/meme media, 3.) related to COVID skepticism/denial, anti-vaccine ideology, and/or conspiracism, and 4.) discursive significance, based on quantitative and qualitative evaluation.

The five corpora coded in this first round included: the *Plandemic* propaganda documentary, the *Vaxxed* propaganda documentary, official posts from the Facebook group of the anti-vaccine and COVID-skeptic organization The Children’s Health Defense (going back three months, excluding comments), the “fake news” blog *Off-Guardian*’s “COVID Factchecker” vertical (comprised of seven blog posts), and the Instagram account “COVID Funny Memes” (constituting 100 pieces of relevant content, excluding comments). The discursive significance of these channels was established based on the following criteria: the films *Vaxxed* and *Plandemic* enjoyed widespread distribution on online platforms (Duchsherer et al., 2020; Kearney et al., 2020); the Children’s Health Defense organization’s Facebook page has over 145,000 followers and is headed by celebrity vaccine detractor Robert F. Kennedy Jr. (Bradshaw et al., 2020); the “Funny COVID Memes” page had only approximately 5,000 followers, but was selected for its apparent ideological neutrality (i.e. both vaccine-denialist and pro-vaccine content appeared in roughly equal proportion); the *Off Guardian* blog represented the least widely consumed inclusion, ranking 74,278 in global internet engagement according to *Alexa* rankings—it was chosen in order to include a text-only coding source and due to its clear COVID/vaccine skeptic position.

Sampling for the second round was likewise purposive, with a focus on capturing media and audiences that might have been missed during Round 1. Channels that spoke to minority demographics were specifically sought out. Once again, five channels were selected for coding based on the same criteria as Round 1. The five corpora constituting the second round of coding were: the Twitter account of Joyce Brooks (a representative of the Denver NAACP and vaccine denialist), the Twitter account of Toby Rogers (a popular and prolific anti-vaccine figure with a following of 29.7k at the time of sampling), the YouTube video *A Message to Aussie Muslims* by Sufyaan Khalifa (urging resistance to COVID public health measures), the *Gardasil Girls* “vaccine injury” Instagram account (including top comments), and the Vaccines Uncovered Instagram account. For all four non-video accounts, 50 individual posts, including retweets and reposts were collected.

The purpose of this selection methodology was not to select the most popular or influential COVID/vaccine-skeptic/denialist channels, but to select a spread of channels which could reasonably be expected to move the research team toward theoretical saturation. Saturation was defined by the research team as that point at which “additional data do not lead to any new emergent themes” (Given, 2015, p. 135).

### Coding methodology

Round 1 corpora were loaded into NVivo, and each member of the four-person codebook team independently coded each corpora. Codes were then compared and consolidated. Infrequent codes were set aside. In the end, preliminary coding produced 34 unique codes pertaining to the narrative tropes and rhetorical strategies which circulate in the anti-vaccine and COVID-skeptic media.

These initial 34 codes were then applied to the second round of corpora. Round 2 corpora were coded according to the preliminary codebook produced in Round 1, with new codes added as deemed appropriate by the coding team. The coding schema developed by each individual member of the codebook team were compared and consolidated. An additional 29 codes were identified during the second round of coding. However, these codes were either found only sporadically or could be consolidated into existing codes. These codes were then analyzed according to an action research methodology approach (Koshy, 2011). The rates of code appearance across corpora were analyzed in comparison with their frequency within corpora. Based on repetition of themes, it was determined that codes pertaining to general audiences (i.e. not demographically specific) had reached a point of adequate saturation. However, in keeping with action research methodology, some relatively infrequent codes were kept. This was done in order to to address codes specific to minority demographics.

The codebook team drafted detailed explanations of the codes and then grouped them according to whether they referred to a narrative trope or a rhetorical strategy, based on the criteria outlined in the literature review. Narrative tropes were further organized according to the implicit antagonist of the narrative. Implicit antagonists were determined based on close reading of content from which these codes were derived. Rhetorical strategies which frequently occurred alongside specific narrative tropes were noted. Examples from the corpora were assigned to each code in order to further illustrate.

## Findings: Rhetorical Strategies and Antagonists

The results of our coding show that online, English-language, anti-vaccine and COVID-denial content can be classified according to twenty-two narrative tropes and sixteen rhetorical strategies (see appendix Tables 1&2). The twenty-two narrative tropes are, in turn, directed toward four major types of antagonists: the government, the medical establishment and political/economic elites; mainstream (and implicitly, pro-vaccine) society at large; an entirely unspecified “shadowy villain”; and the vaccine itself. Some additional unclassified antagonists form a miscellaneous category (see Tables 1 and 2). The ten most frequent codes (“Vaccine Injury,” “Sinister Origins,” Freedom Under Siege,” “Health Freedom,” “Think of the Children!” “Do Your Own Research,” “Heroes and Freedom Fighters,” and “Panic Button) appeared across all four major platforms from which data was sampled: Youtube, Twitter, Facebook, and Instagram. However, it should be noted that salience cannot be determined solely by the frequency with which a code appeared either within or across channels. In keeping with this study’s action research methodology, other factors, such as interaction with other narratives and rhetorics and subjective judgements of affective intensity, were taken into account when determining salience.

**Table 1:**
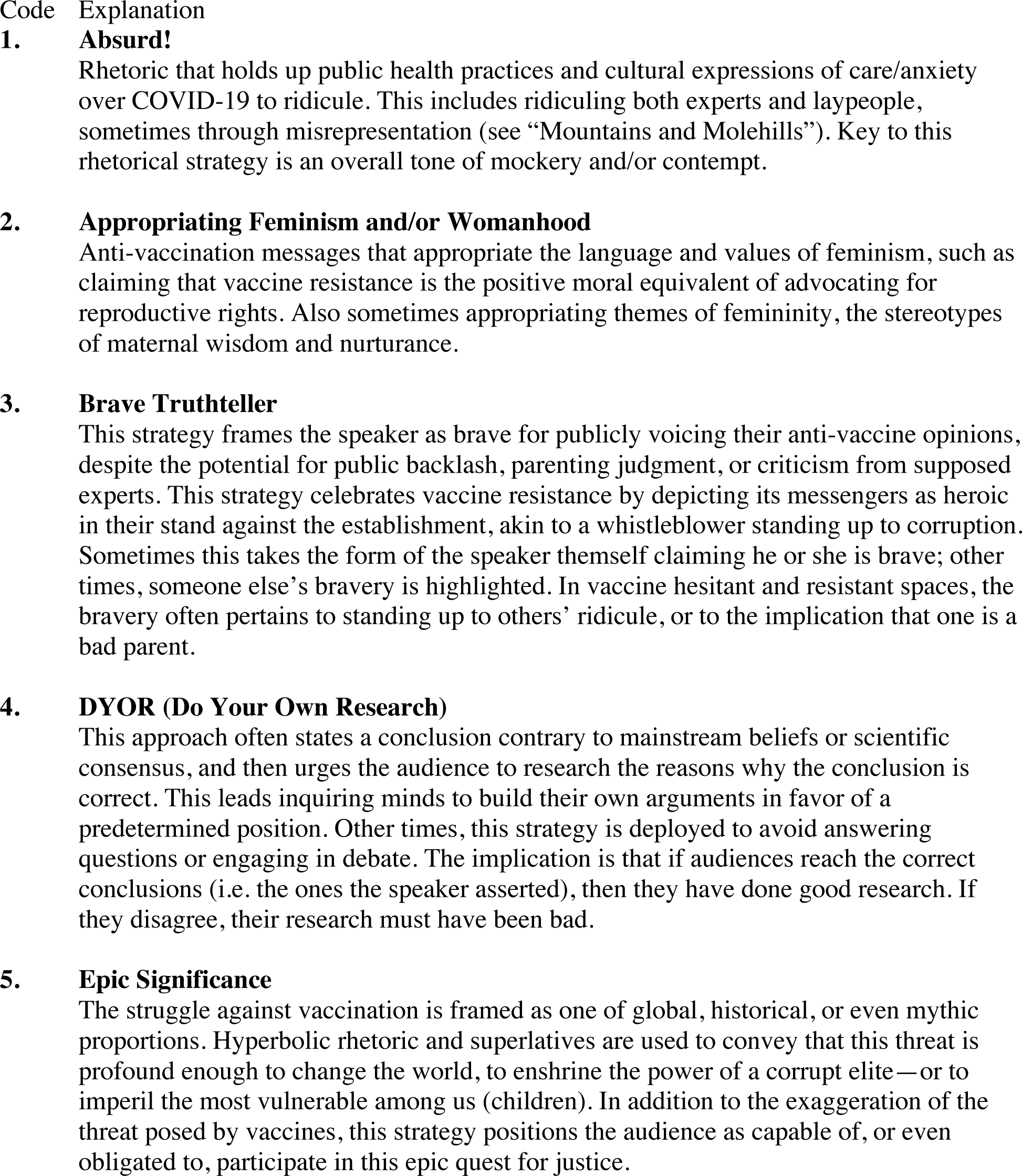
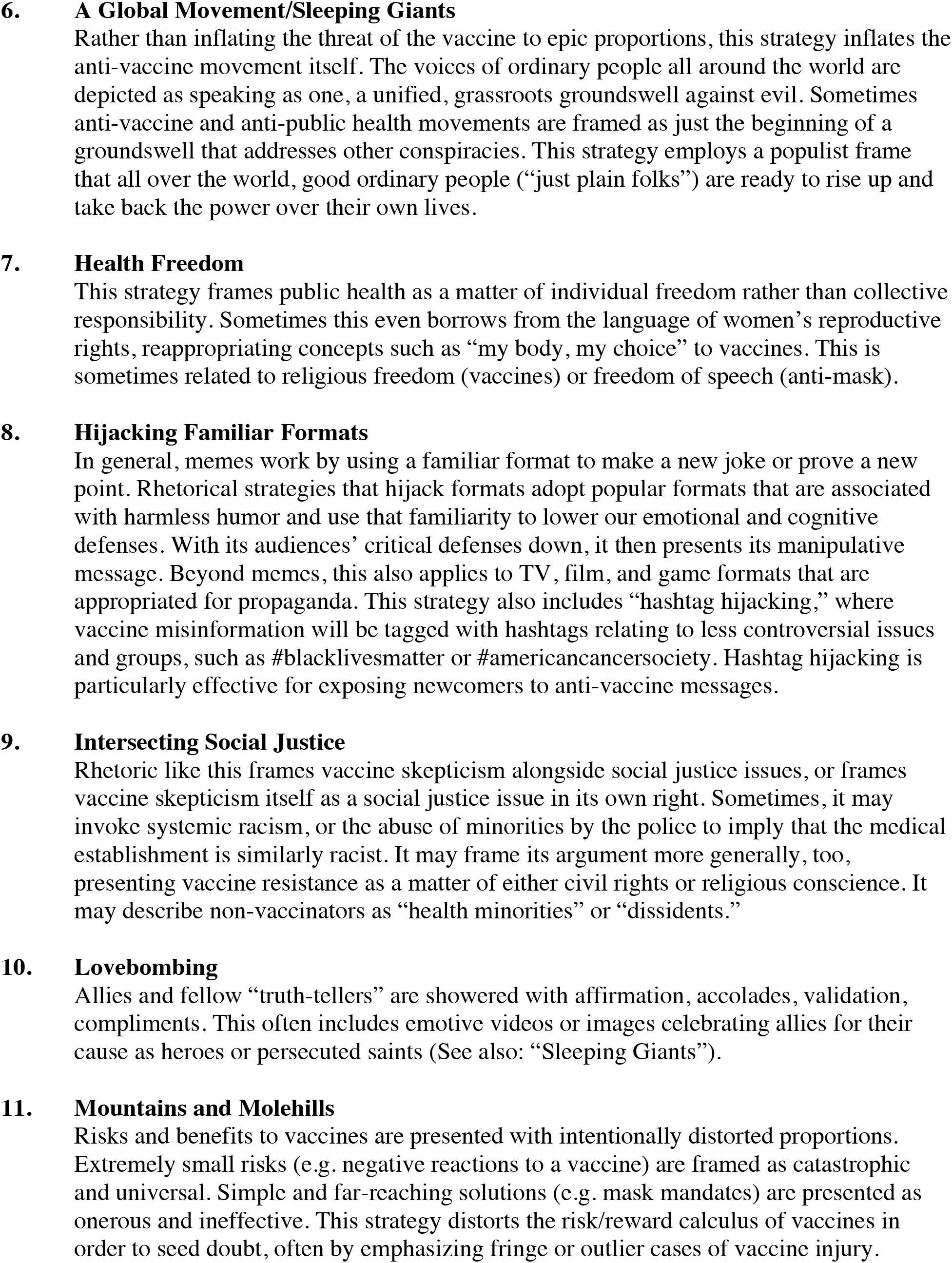
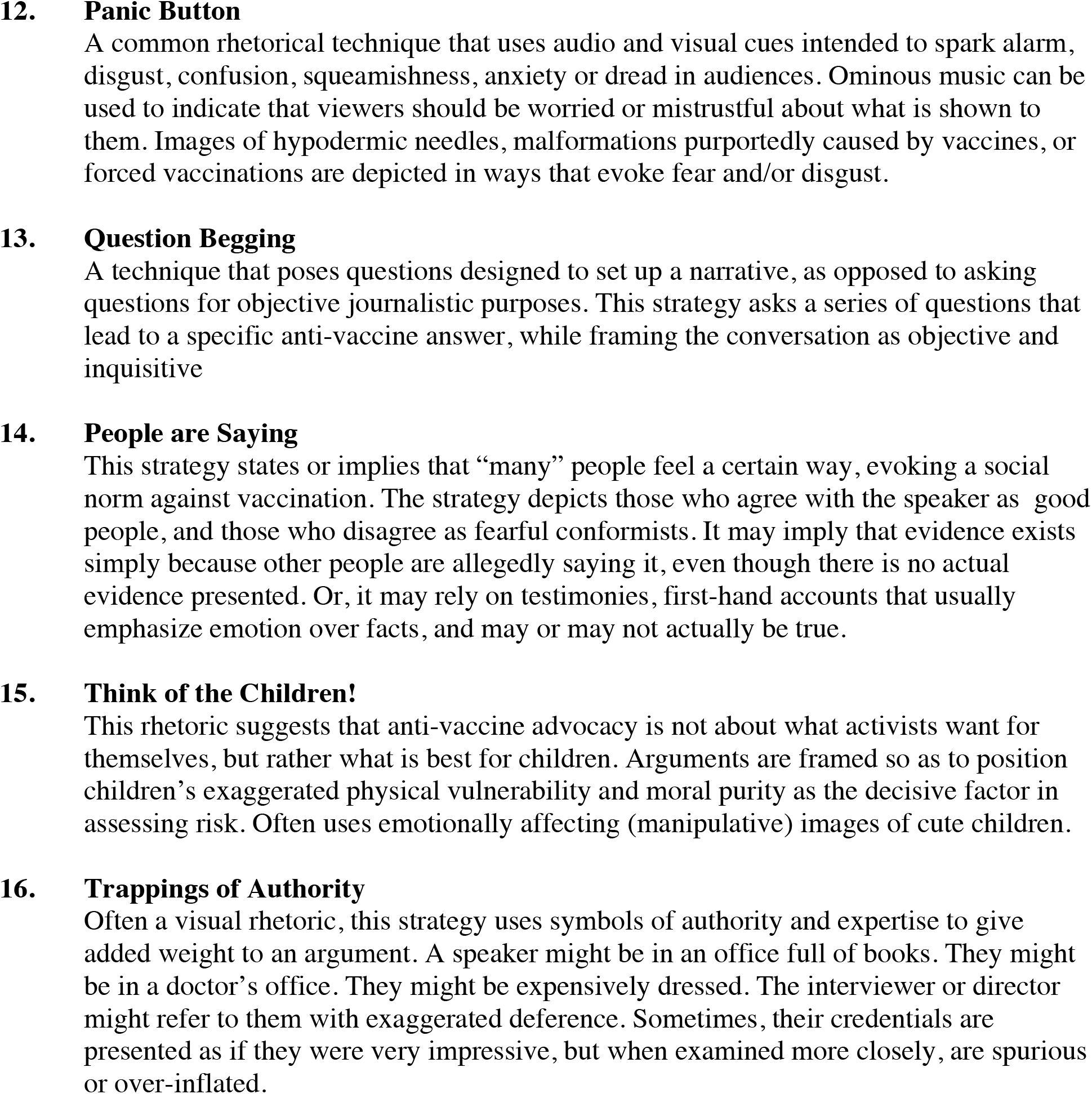
Rhetorical Strategies. *An appendix of visual examples drawn from the dataset is available upon request from*

**Table 2:**
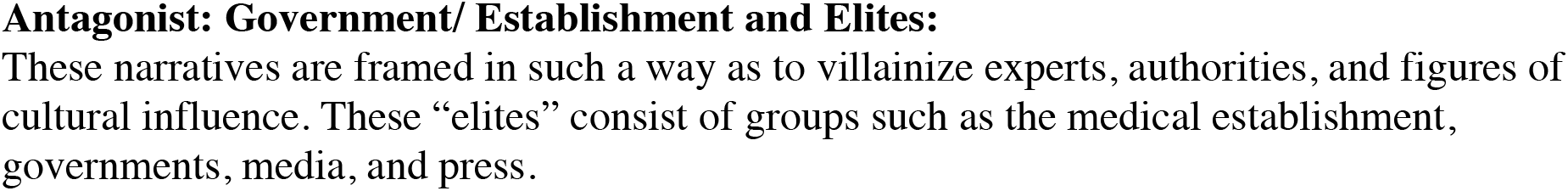
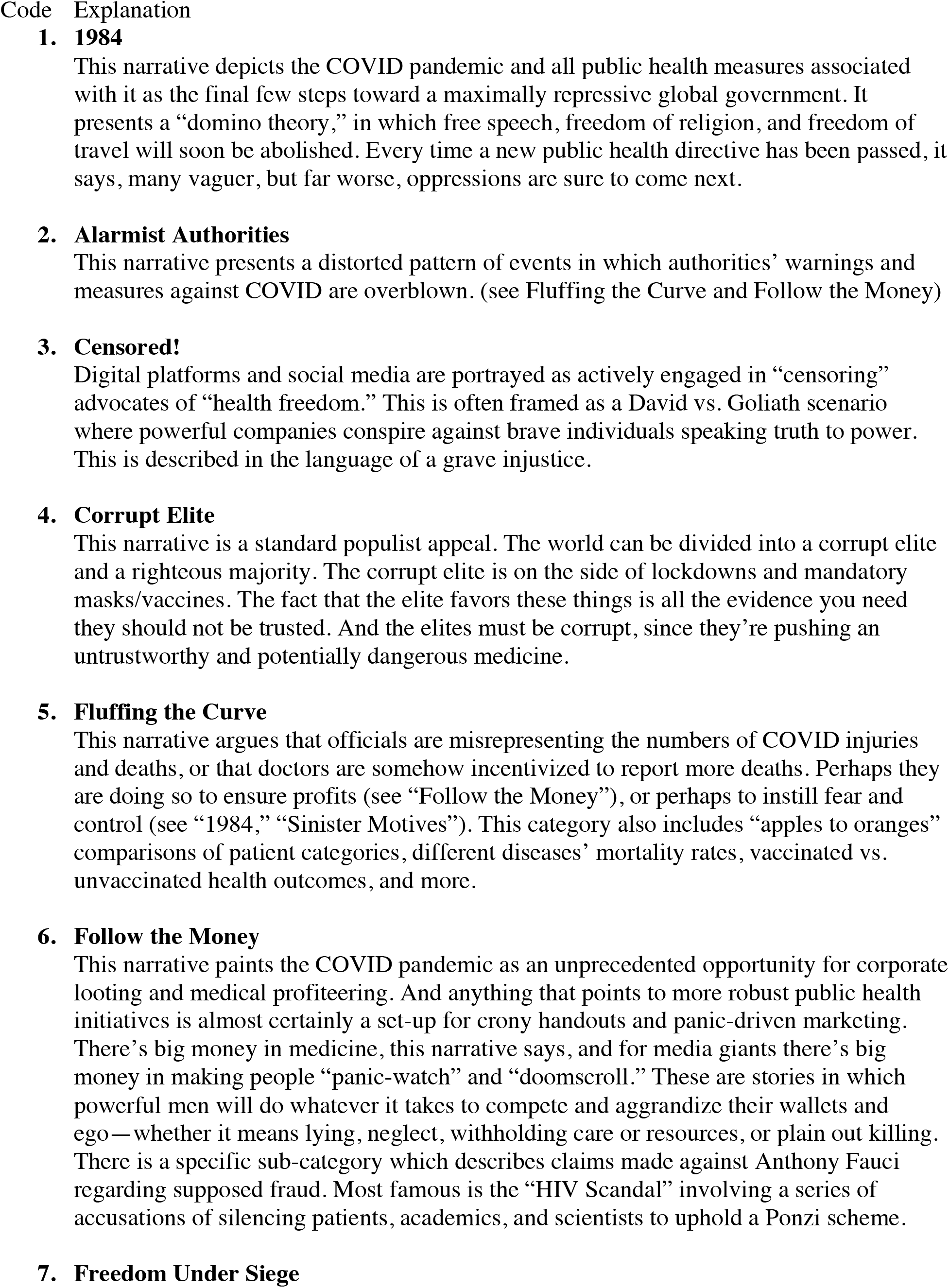
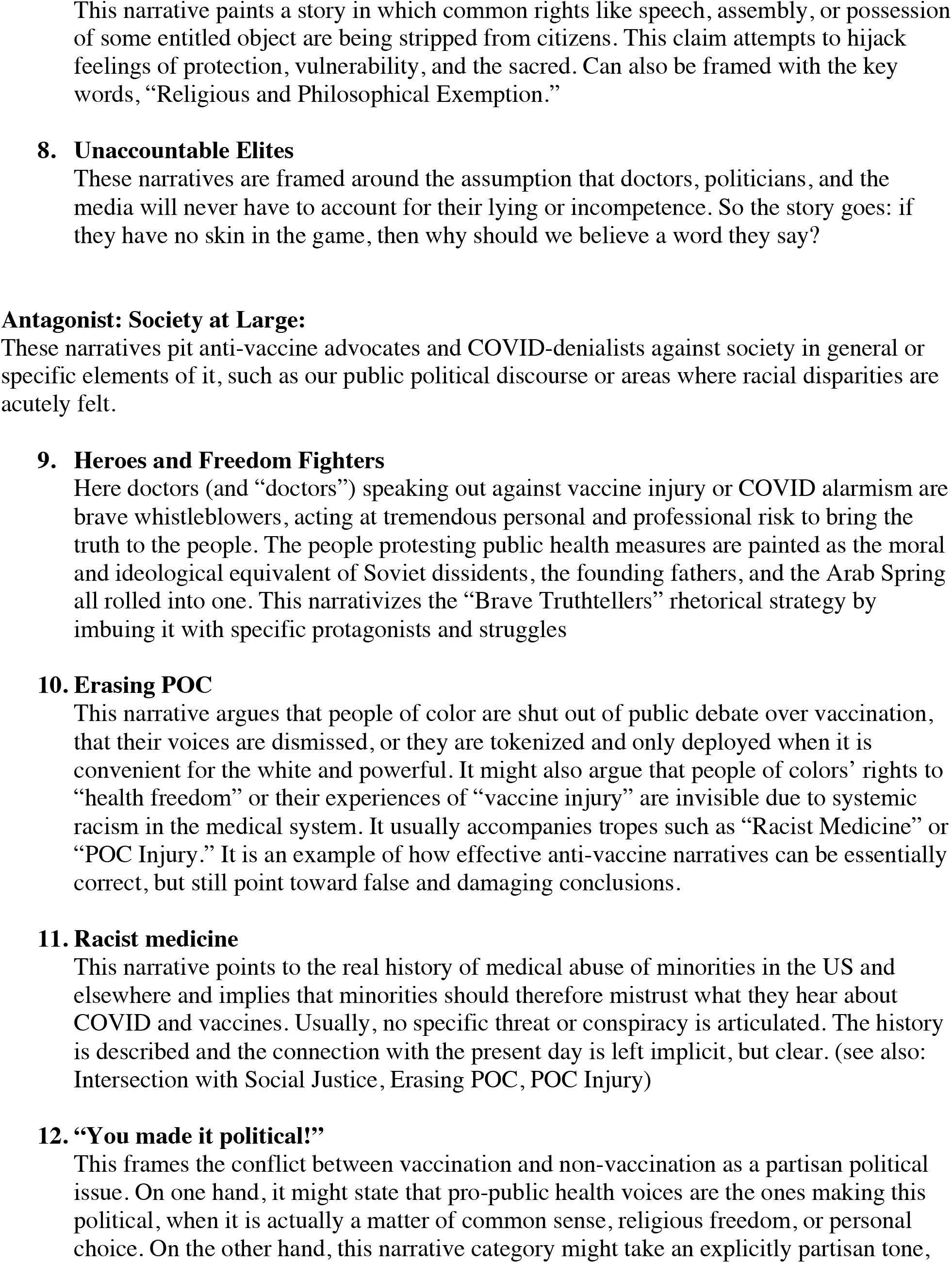
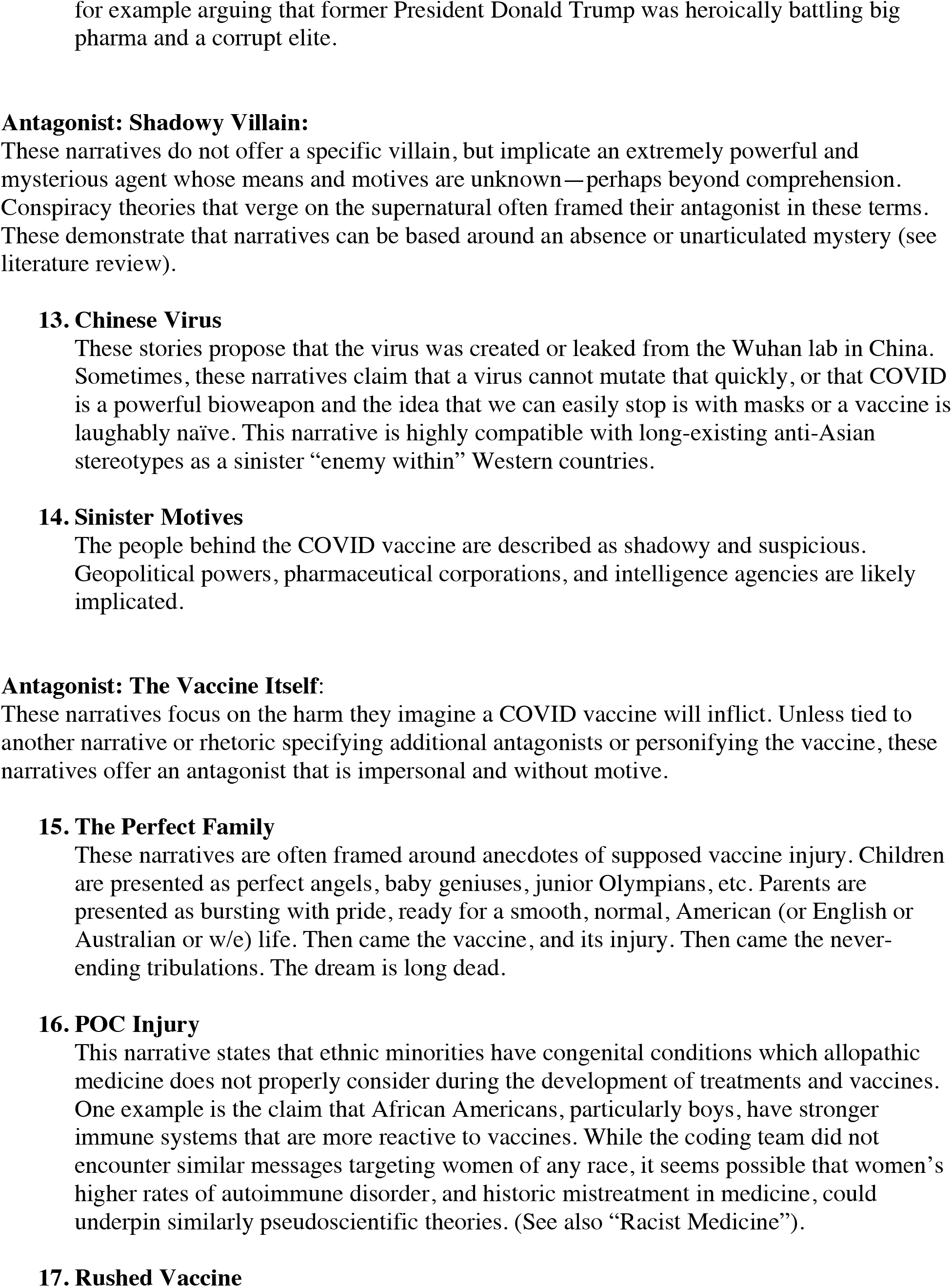
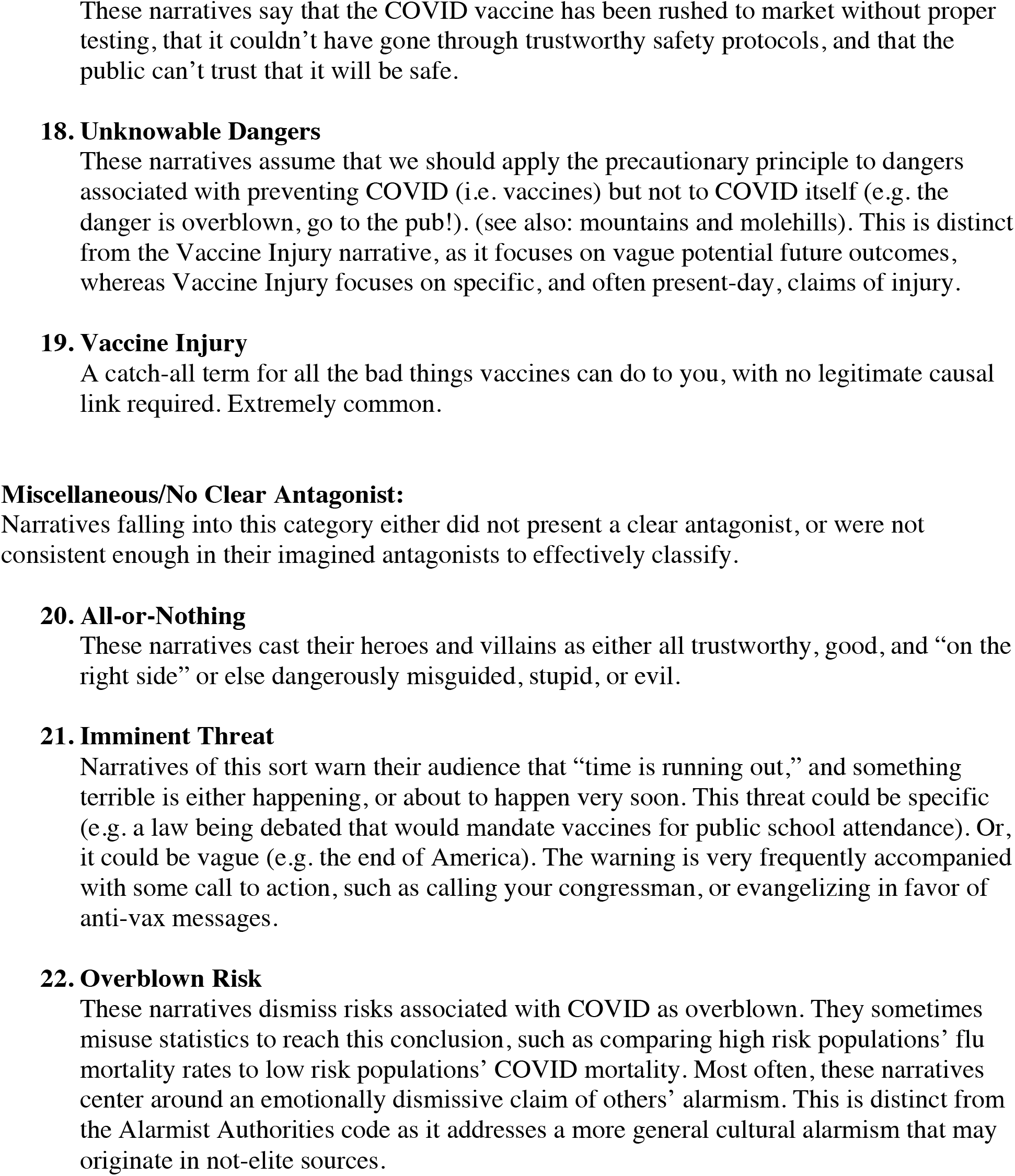
Narrative Tropes (organized by primary antagonist) *An appendix of visual examples drawn from the dataset is available upon request from*

**Table 3:**
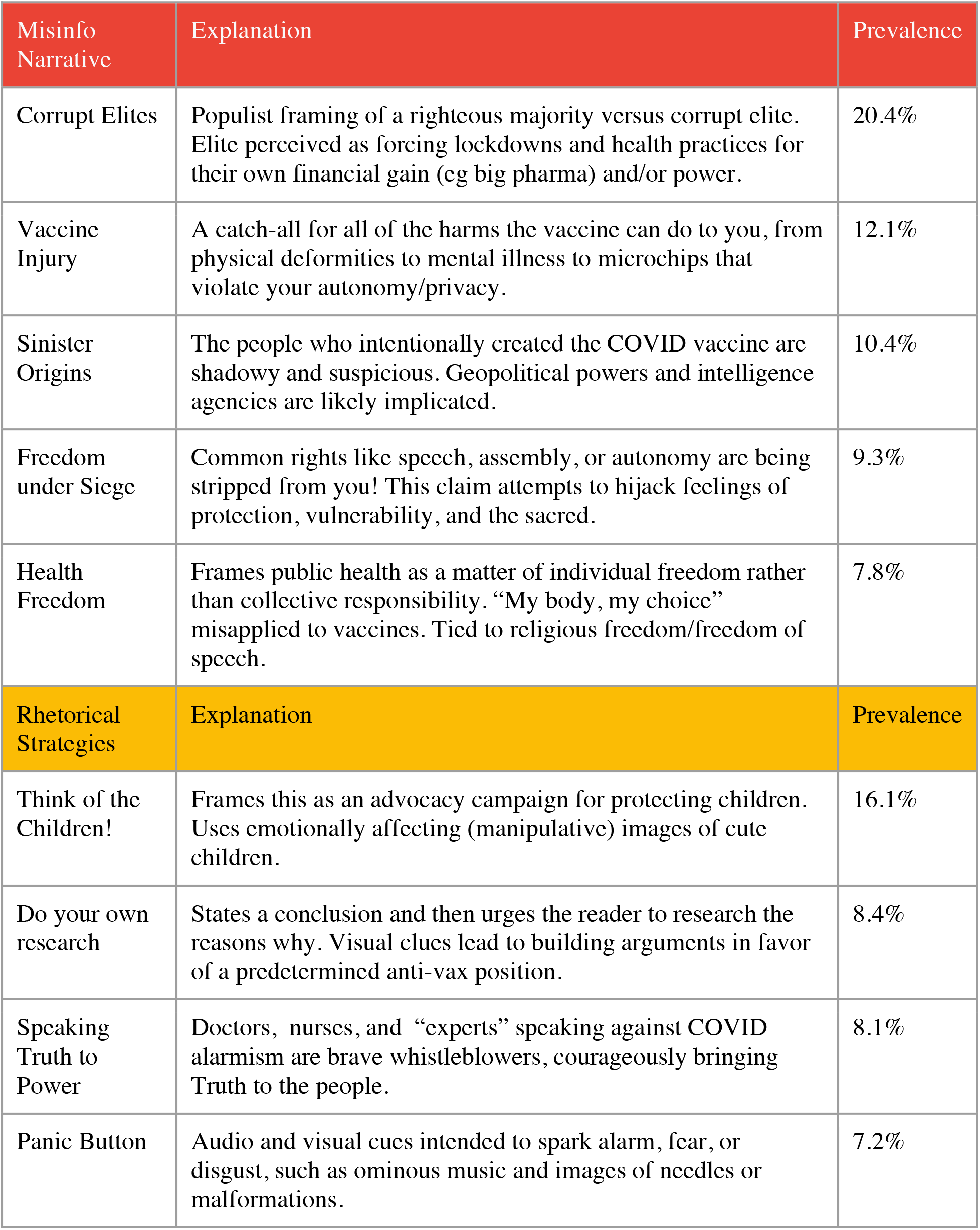
Table of top 10 codes by prevalence.

| Misinfo Narrative | Explanation | Prevalence |
| --- | --- | --- |
| Corrupt Elites | Populist framing of a righteous majority versus corrupt elite. Elite perceived as forcing lockdowns and health practices for their own financial gain (eg big pharma) and/or power. | 20.4% |
| Vaccine Injury | A catch-all for all of the harms the vaccine can do to you, from physical deformities to mental illness to microchips that violate your autonomy/privacy. | 12.1% |
| Sinister Origins | The people who intentionally created the COVID vaccine are shadowy and suspicious. Geopolitical powers and intelligence agencies are likely implicated. | 10.4% |
| Freedom under Siege | Common rights like speech, assembly, or autonomy are being stripped from you! This claim attempts to hijack feelings of protection, vulnerability, and the sacred. | 9.3% |
| Health Freedom | Frames public health as a matter of individual freedom rather than collective responsibility. “My body, my choice” misapplied to vaccines. Tied to religious freedom/freedom of speech. | 7.8% |
| Rhetorical Strategies | Explanation | Prevalence |
| Think of the Children! | Frames this as an advocacy campaign for protecting children. Uses emotionally affecting (manipulative) images of cute children. | 16.1% |
| Do your own research | States a conclusion and then urges the reader to research the reasons why. Visual clues lead to building arguments in favor of a predetermined anti-vax position. | 8.4% |
| Speaking Truth to Power | Doctors, nurses, and “experts” speaking against COVID alarmism are brave whistleblowers, courageously bringing Truth to the people. | 8.1% |
| Panic Button | Audio and visual cues intended to spark alarm, fear, or disgust, such as ominous music and images of needles or malformations. | 7.2% |

### Narrative Tropes

Of the twenty-two narrative tropes coded, four key common narrative tropes were: “Vaccine Injury,” “Corrupt Elite,” “Heroes and Freedom Fighters,” and “Sinister Motives.” Each of these narratives addressed a separate antagonist, representing all categories of antagonist—the Government/The Medical Establish/Media, Society at Large, a Shadowy or Unknown Villain, and the Vaccine Itself—except for “Misc. / No Clear Antagonist.”

The narrative of “Corrupt Elite” was the most frequently coded narrative. This offered a standard populist appeal in which an innocent put disempowered “silent majority” suffer under the tyranny of a powerful and corrupt minority (Mudde 2004; Miller-Idriss 2019). In a process of circular logic, the fact that the government, medical establishment, and high profile cultural figures support such measures as lockdowns, masks, and vaccines is taken as sufficient evidence that these measures are untrustworthy. The narrative of a corrupt elite was very frequently seen with related but distinct codes. “Follow the Money,” for example, was often co-occuring, offering financial motives for elite corruption. “Sinister Motives” offers malevolent and even supernatural explanations for elite missteps. The “Unaccountable Elites” narrative frames government and the medical establishment as immune to consequences from the bad or incompetent actions; it is motive agnostic, but argues that whatever their motives, elites bear no consequences for the mistakes and therefore are always to be mistrusted and disobeyed. These, along with other compatible narratives, point to a larger metanarrative that pits potential vaccine recipients against all of the social institutions working to provide them with a vaccine.

“Vaccine Injury” appeared the most salient of narratives, though not the most frequent. The code was as a catch-all concept describing all negative outcomes (almost exclusively imagined or untrue) associated with taking a vaccine. Examples of this narrative rarely provided a clear causal link between vaccination and injury. Instead, it most frequently juxtaposed a claim to having been vaccinated alongside a claim to injury. These injuries were often vaguely described and only infrequently accompanied by claims to an actual diagnosis of malady. Messages conveying this narrative frequently used the “Panic Button” style of audio-visual rhetoric, in which images of needles, crying infants, unsettling sounds or unattractive colors are employed to produce feelings of disgust and unease.

A third popular narrative, “Heroes and Freedom Fighters” is compatible with populist framings, presenting medical doctors (as well as chiropractors and naturopaths) as brave whistleblowers, risking their reputations and careers by speaking truth to power. This narrative often coincided with rhetorics of “Brave Truthteller,” “Sleeping Giants,” and “Health Freedom,” presenting the people organizing against public health measures as the moral equivalent of pro-democracy dissidents.

### Rhetorical Strategies

Of the sixteen rhetorical strategies coded, four key, common rhetorical strategies were: the “Brave Truthteller,” “Do-Your-Own-Research (DYOR),” “Mountains and Molehills,” and “A Global Movement/Sleeping Giants”. Each of these rhetorical strategies was present across multiple categories of narrative antagonist. For example: the rhetorical strategy labeled “Brave Truthteller”—in which a speaker claims to be speaking a dangerous truth which “the establishment” or society at large is suppressing—was significantly present in all four categories of antagonists and even in those relatively few narratives with no clear antagonist.

Another common rhetorical strategy that is directed toward multiple protagonists is “DYOR” or “Do-Your-Own-Research.” DYOR works by trying to empower the audience to develop their own bodies of evidence and methods of reasoning in order to reach a preordained conclusion. Sometimes, this leads inquiring minds to build their own arguments in favor of a predetermined position. Other times, DYOR offers vaccine denialists and skeptics a means to avoid answering questions or engaging in debate: they may simply demand that the interlocutor should do their own research. If the interlocutor reaches the correct conclusions (i.e. anti-vaxx conclusions), then they’ve done good research. If they still disagree, so the reasoning goes, their research must have been bad.

A third popular category is what we refer to as “Mountains and Molehills.” In this rhetorical strategy, vaccines’ risks and benefits are presented without a proper sense of proportion. Extremely small risks (e.g. negative reactions to vaccines) are framed as catastrophic and universal. Simple and far-reaching solutions (e.g. mask mandates) are presented as onerous and ineffective. The risk/reward calculus is thereby severely distorted. This strategy is used to seed doubt via emphasizing fringe or outlier cases.

Presenting COVID-19 vaccine resistance as a part of a global movement of “sleeping giants”—honest, everyday citizens who are on the cusp of rising up against an oppressive “global elite”—was a common rhetorical strategy that cut across narratives targeting all categories of antagonists. This rhetoric frames its accompanying narrative to suggest that the voices of ordinary people all around the world are articulating that narrative as a unified mass. Anti-vax and anti-public health movements are presented to be just the beginning of this groundswell. All over the world, so the trope goes, self-conceived “ordinary people” are ready to rise up and take back the power over their own lives. This rhetoric taps into the populist persuasive strategies and schemata outlined above, which frame the pure, ordinary people against the nefarious, evil elite. In some cases, populist rhetoric intersects with nationalist rhetoric, arguing that only a stronger state can save ordinary people from bad elites. In other cases, these populist frames intersect with anti-government resistance, framing elites as not only out-of-touch with the needs of the pure, ordinary people but actively working against them in tyrannical ways that warrant uprisings, revolution, armed resistance or even a new civil war.

## Discussion

The scope of this study is intended to create a codebook of online anti vaccination narratives and rhetoric, so to support government officials engaged in managing disinformation during the COVID-19 vaccination campaign. Local, national, and international governments, as well as civil society organizations, need to be prepared to manage the infodemic by promoting the timely dissemination of accurate information based on science and evidence, in particular high-risk groups. This dissemination of accurate information should be both positive and defensive. That is, messaging campaigns must communicate both the latest in scientific understanding of COVID, its spread and prevention, and effective counter-messages against misinformation. The codes identified by this study offer governments and civil society groups a catalogue of anti-vaccine message styles that may assist in the latter efforts.

Pro-vaccine audiences, the vaccine hesitant, and the wholly agnostic can generally be addressed as a single group. Anti-vaccine belief holders, a much smaller subset of the overall vaccine hesitancy spectrum (SAGE Working Group, 2014), however, must have counter-messaging tailored specifically for them. This comports with the distinction between “prophylactic” and “therapeutic” counter-messaging (Compton, 2020). Evidence demonstrates that prophylactic exposure to counter-messaging would prompt hesitant, agnostic, and pro-vaccine audiences to develop their own counterarguments against misinformation and disinformation (Banas & Rains, 2010; Pfau et al., 2005). Furthermore, these audiences would not need to be warned against every type of misinformation or disinformation they might encounter. Exposure to counter-messaging against one dimension of a mis/disinformation campaign confers resistance to other dimensions associated with that topic of mis/disinformation (Braddock, 2020; McGuier, 1962; Papageorgis & McGuire, 1961). This phenomenon is sometimes called the “blanket of protection” (Ivanov et al., 2016; Parker et al., 2016). So, for example, an effective counter-message addressing exaggerated claims of vaccine injury can in theory be expected to confer some resistance to any other narrative trope or rhetorical strategy listed in this codebook.

The joint effects of targeted counter-messaging and blanket of immunity might best be mobilized through counter-messaging that addresses tropes and/or strategies that appear most frequently, and that are thematically linked to other denialist narratives and rhetoric. For example, the versatility of rhetorical strategies such as “Brave Truthtellers” may be both a cause and a symptom of their effectiveness; its versatility lends itself to a variety of narratives, while its repetition across narratives enhances its effectiveness through repetition. Counter-messaging that alerts audiences to the manipulative persuasion of the “Brave Truthteller” strategy will therefore be effective against a variety of misinformation and disinformation utilizing the strategy, as well as likely conferring further future resistance against the rhetoric and narratives that appear alongside it.

Similarly, “Do Your Own Research” is also a popular rhetorical strategy throughout conspiracy cultures, like QAnon. QAnon disinformation networks have been shown to amplify anti-vaccination rhetoric and messaging (Timberg & Dwoskin, 2021), which raises important questions about the impact of overlapping persuasive rhetorical strategies in addition to amplification through hashtags or coordinated campaigns. By counter-messaging against transnetwork themes, a blanket of immunity may protect audiences from anti-vaccine and COVID-denialist themes that are not described in this codebook or have yet to emerge.

Counter-messaging to address audiences who already hold undesirable viewpoints (i.e. therapeutic counter-messaging) is a newer and less well-established process than its prophylactic counterpart (Compton, 2020; Ivanov et al., 2016; M. L. M. Wood, 2007). A potential for backlash is always present in counter-messaging campaigns (Lee, 2019; Matland & Murray, 2013). And while the so-called “backlash effect” (that is, the theory that factual counterargument entrenches false beliefs) has been credibly challenged (T. Wood et al., 2019), there is ample evidence that carelessly repeating false information can help spread it (De keersmaecker et al., 2020; Delouvée, 2020; Fazio et al., 2015; Skurnik et al., 2005). Therefore, public health messaging that addresses anti-vaccination audiences, already hardened in their beliefs, must be preceded by especially rigorous testing. This codebook is presently informing a series of tests to determine the efficacy of public messaging that addresses some of the narratives and rhetoric found in it. The results of that study are expected to be available in pre-print by early summer 2021.

## Conclusion

This paper reports on the qualitative classification of online, English-language anti-vaccination rhetoric about a COVID-19 vaccine. Our analysis of sixteen persuasive rhetorical strategies and twenty-two anti-vaccination messages directed toward specific antagonists is necessarily descriptive at this stage. We hope this work inspires additional empirical research to help illuminate the following questions: which of these rhetorical strategies and messages are most encountered online and for which demographic groups? Which of the messages carries the most persuasive appeal? How can persuasive rhetorical strategies be effectively countered in online spaces? How might different narratives and rhetorics appeal to audiences in different countries, different demographics and subcultures, and in different languages? Given the rise in populist movements across many countries, it seems possible that similar antagonisms would be articulated in anti-vaccine and COVID-denialist media content. However, given the highly contextual and referential qualities of memes and other social media content, these antagonisms could reasonably be expected to assume significantly different expressive forms. To avoid the risk of blowback, it is essential that similar coding studies and counter-message testing be undertaken prior to the launch of public health campaigns addressing anti-vaccine and COVID-denialist mis- and disinformation.

## Data Availability

Data referred to in manuscript are available. Please contact.

## Notes

### Competing Interest Statement

The authors have declared no competing interest.

### Funding Statement

Funding for this study was provided by Jigsaw, Google LLC, New York, NY

### Author Declarations

American University Institutional Review Board

